# Towards Understanding Autism Heterogeneity: Identifying Clinical Subgroups and Neuroanatomical Deviations

**DOI:** 10.1101/2023.10.21.23297299

**Authors:** Jente Meijer, Bruno Hebling Vieira, Camille Elleaume, Zofia Baranczuk-Turska, Nicolas Langer, Dorothea L. Floris

## Abstract

Autism Spectrum Disorder (‘autism’) is a neurodevelopmental condition characterized by substantial behavioural and neuroanatomical heterogeneity. This poses significant challenges to understanding its neurobiological mechanisms and developing effective interventions. Identifying phenotypically more homogeneous subgroups and shifting the focus from average group differences to individuals is a promising approach to addressing heterogeneity. In the present study, we aimed to parse clinical and neuroanatomical heterogeneity in autism by combining clustering of clinical features with normative modeling based on neuroanatomical measures (cortical thickness [CT] and subcortical volume) within the ABIDE datasets (N autism=861, N neurotypical=886, age-range 5-64). First, model-based clustering was applied to autistic symptoms as measured by the Autism Diagnostic Observation Schedule to identify clinical subgroups of autism. Next, we examined whether clinical subgrouping resulted in increased neurobiological homogeneity within the subgroups compared to the entire autism group by comparing their spatial overlap of neuroanatomical deviations. We further investigated whether the identified subgroups improved the accuracy of autism classification based on the neuroanatomical deviations using supervised machine learning with cross-validation. Results yielded two clinical subgroups primarily differing in restrictive and repetitive behaviours (RRB). Both subgroups showed increased homogeneity in localized deviations with the high-RRB subgroup showing increased volume deviations in cerebellum and the low-RRB subgroup showing decreased CT deviations predominantly in the postcentral gyrus and fusiform cortex. Nevertheless, substantial within-group heterogeneity remained highlighted by the failure of the classifier to distinguish between the subgroups. Identifying subgroups of autism has substantial clinical implications opening the potential for more tailored behavioural interventions and improving clinical outcomes.

**General Scientific Summary:** Autism is characterized by pronounced behavioural and neurobiological heterogeneity. This study suggests that reducing this heterogeneity at the clinical level by employing subgrouping also results in more similar neuroanatomical profiles in the identified clinical subgroups. Although we could demonstrate that clinical subgrouping reduces neuroanatomical heterogeneity, simultaneously, there remained substantial heterogeneity potentially pointing towards multiple pathways resulting in high RRBs and neurobiological subgroups with similar clinical phenotypes.

## Introduction

Autism is a lifelong, heterogeneous neurodevelopmental condition characterized by differences in social communication and restricted repetitive behaviours and interests (RRBs; American Psychiatric Association, 2013). Prevalence estimates have risen significantly in recent years, suggesting that 1 in 36 children has autism (Maenner et al., 2021). While this can incur a considerable economic cost for society (Buescher et al., 2014), at the individual level, the condition can also have a profound impact on the quality of life of the autistic person (Moyal et al., 2014; Oakley et al., 2021) who may require substantial medical and behavioural support. Still, most interventions have limited efficacy (Bacon et al., 2014) as they are not optimally tailored to each individual’s clinical and neurobiological phenotype. Reducing this pronounced heterogeneity both at the clinical and neurobiological level is among the biggest current challenges to pave the way for more individualized support services for those who want and require them.

An increasingly popular strategy for addressing clinical heterogeneity involves the delineation of subgroups to foster greater group homogeneity (Lai et al., 2013). The identification of clinically meaningful subgroups is crucial for targeted support, particularly considering the current prominence of behavioural programmes as the most efficacious mode of intervention (Baribeau et al., 2022). First efforts have employed clustering algorithms to identify subgroups with different clinical feature compositions (Bitsika et al., 2008; Cholemkery et al., 2016; Georgiades et al., 2013; Hu & Steinberg, 2009; Hus et al., 2007; Lefort-Besnard et al., 2020; Lombardo et al., 2016; Mandelli et al., 2023). These studies have utilized clinical measures, such core clinical features (e.g., Autism Diagnostic Observation Schedule [ADOS], Autism Diagnostic Interview [ADI]), adaptive functioning or higher-order cognitive domains to identify between three to five subgroups which were characterized by different symptom profiles. However, among these, some studies show that identified subgroups may most strongly differ by demographic variables such as age, full-scale IQ (FIQ) and sex (Bitsika et al., 2008; Lefort-Besnard et al., 2020). These intertwined associations between these subgroups and demographic variables introduce ambiguity as to whether the observed subgroup variations stem from clinically relevant metrics or basic demographic measures. Furthermore, the studies investigating autism subgroups did not further examine whether these clinically differentiated subgroups might also yield heightened homogeneity in brain structures. Such homogeneity in brain structures could potentially provide insights into the specific neuroanatomical foundations underlying distinct autistic traits.

The neurobiological underpinnings of autism have been extensively studied, particularly in terms of cortical thickness (CT) and volume. While most studies show differences in brain structure between autistic individuals and non-autistic individuals (NAI), findings are predominantly inconsistent with some studies reporting thicker cortex, thinner cortex or no significant differences in autistic individuals (Duerden et al., 2014; Durkut et al., 2022; Ecker et al., 2022; Hazlett et al., 2011; Khundrakpam et al., 2017; Mensen et al., 2017; Pretzsch et al., 2022; Schaer et al., 2013; Shen et al., 2022; Wallace et al., 2015; Zielinski et al., 2014). This well-documented heterogeneity in autism may contribute to this inconsistency in findings. There is thus a need for adopting novel methods and shifting away from case-control paradigms that assume that the group mean is representative of the entire population, which may not be accurate for heterogeneous populations like autism (Marquand et al., 2016, 2019; Rutherford et al., 2022, 2023).

A promising novel method for addressing the neurobiological heterogeneity in mental health disorders is normative modeling (Marquand et al., 2016; Rutherford et al., 2022). A normative model captures the trajectory of the typical developing brain across relevant variables such as age, which can be used to predict CT or volume for each individual and thereby show the deviation from the typical pattern for each individual. Previous studies used normative modeling to identify individualised deviations of CT among autistic individuals, revealing heterogeneous widespread patterns of deviations (Bethlehem et al., 2018, 2020; Segal et al., 2023; Zabihi et al., 2019, 2020). Clustering at the neurobiological level has further been used on the basis of deviations derived from normative models (Bethlehem et al., 2020; Shan et al., 2022; Zabihi et al., 2020). Clustering at the clinical level in combination with normative models has been used in predefined language subgroups only (Floris et al., 2021). However, no previous studies have used normative-model derived neuroanatomical measures to characterize data-driven clinical subgroups.

In the present study, we aimed to reduce clinical and neuroanatomical heterogeneity in autism by identifying clinically distinct subgroups within autism and characterizing their neuroanatomical profile. For this, we leveraged the large-scale, publicly available Autism Brain Imaging Data Exchange (ABIDE) datasets (Di Martino et al., 2014, 2017). We hypothesised to find clinically distinct subgroups showing more consistent brain atypicality. We thus expected a higher homogeneity in CT and volume deviations in the subgroups compared to the entire autism group.

## Methods

### Participants

We combined the Autism Brain Imaging Data Exchange (ABIDE) repositories I (Di Martino et al., 2014) and II (Di Martino et al., 2017). ABIDE I includes in total 1112 participants (539 autism, 573 TC) from 17 different sites, and ABIDE II includes 1114 participants (521 autism, 593 TC) from 19 different sites. Detailed information on imaging acquisition parameters can be found on the ABIDE website (https://fcon_1000.projects.nitrc.org/indi/abide/).

We excluded females due to scarce availability (N autism=130, N NAI=280) and autistic individuals whose ADOS-Generic (ADOS-G) had not been administered by research-reliable personnel (N autism=69). For the clustering analysis (based on clinical data, i.e., ADOS-G), we further excluded participants with missing ADOS-G scores (N autism=348) and FIQ (N=55). This resulted in a sample of 499 autistic individuals who were included in the clustering analysis. For the normative modeling analysis (based on neuroimaging data), we applied the following exclusion criteria: *a)* participants with lacking neuroimaging data (N autism=71, N NAI=37); b) low image quality (N autism=15, N NAI=7); *c)* corrupted preprocessing (N autism=4, N NAI=1); *d)* participants missing FIQ information (N autism=47, N NAI=66); and *e)* sites with less than ten individuals per diagnostic group (Bayer et al., 2022; Gaiser et al., 2023) (N autism=69, N NAI=15). For further details see the SI. This resulted in a sample of 690 autistic and 744 NAI between 5-64 years who were included in the normative modeling analysis (see Table 1). A flowchart depicting the selection process for each analytical step is visualized in Figure S1.

**Table 1.** Participant demographics. The sample size reflects the total number of individuals before applying analyses-specific exclusion criteria for clustering and normative modeling. Abbreviations: NAI = non-autistic individuals. FIQ = full-scale intelligence quotient. ADOS-G = Autism Diagnostic Observation Schedule Generic. RRB = restrictive and repetitive behaviours. SD = standard deviation. Differences between autism and NAI in age and FIQ were determined with a Wilcoxon signed rank test.

|  | Autism (N = 861) |  |  | NAI (N = 886) |  |  |  |
| --- | --- | --- | --- | --- | --- | --- | --- |
|  | *Mean* | *SD* | *Range* | *Mean* | *SD* | *Range* | *p-value* |
| **Age** | 16.12 | 8.88 | 5.1 - 64 | 16.58 | 8.96 | 5.9 - 64 | 0.45 |
| **FIQ** | 106.05 | 17.14 | 41 - 149 | 112.9 | 12.43 | 71 - 148 | < 0.001 |

### Model-based clustering

We aimed to identify clinically distinct subgroups in autism using the ADOS-G (Lord et al., 2000) subscores for social interaction, communication, and restricted and repetitive behaviours (RRBs). After z-standardizing the sub-scores, we additionally accounted for confounding variables, including age, site, FIQ, ADOS-G module, and the other two subscores using a General Linear Model (GLM). The resulting residuals were used for the subsequent analyses.

Next, we employed a model-based clustering method (Banerjee & Shan, 2011) to identify clinical subgroups in autistic individuals. The density estimation was based on parameterized finite Gaussian mixture models estimated by EM algorithm initialized by hierarchical model-based agglomerative clustering. The optimal number of clusters was determined according to the Bayesian Information Criterion (BIC). To assess the stability of the clustering results, we performed bootstrapping with 10.000 iterations and computed the Jaccard similarity index (JI) (Hennig, 2007). Finally, we tested for differences in subscores between the identified subgroups using Wilcoxon signed-rank tests. To avoid inflation of the Type I error rate (Gao et al., 2022), we additionally used the ‘clusterpval’ package in RStudio with 10.000 draws to determine the significant differences in means between the clusters (Gao et al., 2022; Winter & Hahn, 2022). For further details, see the SI.

### Image processing

We processed the T1-weighted images from the ABIDE datasets using the FreeSurfer software (version 7.3.2) (Fischl, 2012). In brief, the FreeSurfer pipeline consists of intensity normalisation, skull stripping and grey (GM) and white matter (WM) surface segmentation, resulting in a tessellated representation of the GM/WM boundary. The resulting surfaces were corrected for topological abnormalities and registered to a spherical atlas based on individual cortical folding patterns. CT was extracted for each individual using the Destrieux parcellation scheme consisting of 74 regions in each hemisphere (Destrieux et al., 2010). To label subcortical tissue classes, we further applied the volume-based stream. After affine registration with MNI305 space, initial volumetric labeling, B1 bias field intensity correction, a high dimensional nonlinear volumetric alignment to the MNI305 atlas was performed and the volume was labeled (Fischl et al., 2002). For more detailed information, see the SI.

### Normative modeling

Normative modeling is a novel statistical framework that allows to compare individual-level neuroanatomical features (CT and subcortical volume) to a reference population (Marquand et al., 2016, 2019; Rutherford et al., 2022, 2023). This method yields coherent measures of predictive confidence in addition to point estimates, meaning we can predict both the expected neuroanatomical changes and the associated predictive uncertainty for each individual allowing us to quantify the regional deviation of CT and subcortical volume from the neurotypical range.

Here, we performed normative modeling using warped likelihood Bayesian Linear Regression (BLR) (Fraza et al., 2021) model implemented in the PCNtoolkit (version 0.26) in Python (version 3.9.16). We included age, site, FIQ, and quality measures (i.e., the Euler number) as covariates and additional intracranial volume for subcortical volumes. The dataset was split into a training set consisting of 90% of NAI and a test set consisting of all autistic individuals and the remaining 10% of NAI. We ensured that age, FIQ and site were evenly distributed across the training and test sets. A separate normative model was built for each of the 148 CT ROIs and 23 subcortical ROIs independently. Figure S3 depicts an example of a centile map and the model’s trajectory for the left occipital pole. Next, we computed normative probability maps (NPMs) consisting of subject-specific Z-scores, which quantify the deviation of each participant from the normative model in each ROI. To evaluate model performance, we obtained state-of-the-art model evaluation parameters (see SI and Figure S4).

### Characterizing homogeneity and heterogeneity

To further characterize the output from the normative model, NPMs were thresholded at an absolute value of Z>|2.6| (i.e., *P* <0.005) (Floris et al., 2021; Segal et al., 2023; Wolfers et al., 2018, 2020) and we defined extreme positive and extreme negative deviations for each participant. Here, a positive deviation score indicated higher CT (subcortical) values compared to the reference model. To illustrate the heterogeneity in autism, we visualized the individual NPMs for the top ten participants with the highest deviation scores (see Figure S5). To assess whether subgrouping increases biological homogeneity in autism, we generated spatial overlap maps separately for positive and negative deviations using the percentage of individuals that showed an extreme deviation in each ROI. These overlap maps depict regions where participants shared common deviations in each group (see Figure 2). Next, we statistically tested for differences in these overlap maps between the two autistic subgroups and between the entire autism group using group-based permutation tests (Segal et al., 2023).

Further, to test the robustness of the results, we conducted an additional sensitivity analysis by controlling the false discovery rate (FDR) separately for each NPM. For further details see the SI. Finally, we assessed biological separability of the subgroups by applying a Support Vector Machine (SVM). We used the deviation Z-scores derived from the normative model for all ROIs as features and evaluated under five-fold cross-validation whether classifying the subgroups from NAI would outperform classifying the entire autism group from NAI (see SI).

The code used for all analyses and visualisation is available on GitHub: https://github.com/JenteMeijer1/Autism-heterogeneity

## Results

### Clinical Subgrouping

Using model-based clustering based on ADOS-G sub-scores, the BIC identified the EEV model (i.e., the best cluster representation was equal in volume and shape and varies in orientation between clusters), as best-fitting Gaussian model and two as the optimal cluster number (see Figure 1A). This cluster number was stable (JI high-RRB subgroup=0.78; JI low-RRB subgroup=0.92). The clusterpval package showed that the two subgroups significantly differed across the core clinical ADOS-subscores (Euclidean distance = 1.430, *p* = 0.027). Specifically, the two clinical subgroups were primarily distinguished by differences in RRBs (*W*=42752, *p*<0.001) and to a lesser extent by communication scores (*W*=19396, *p*=0.03), as depicted in Figure 1B and Table 2. Specifically, subgroup 1 showed significantly higher RRBs, while subgroup 2 demonstrated significantly higher communication scores (Table 2). In the following, we will refer to subgroup 1 as the high-RRB subgroup and subgroup 2 as the low-RRB subgroup. The subgroups did further not differ in age, FIQ, or ADOS-G module (see Figure S2 and Table 2).

**Figure 1.**
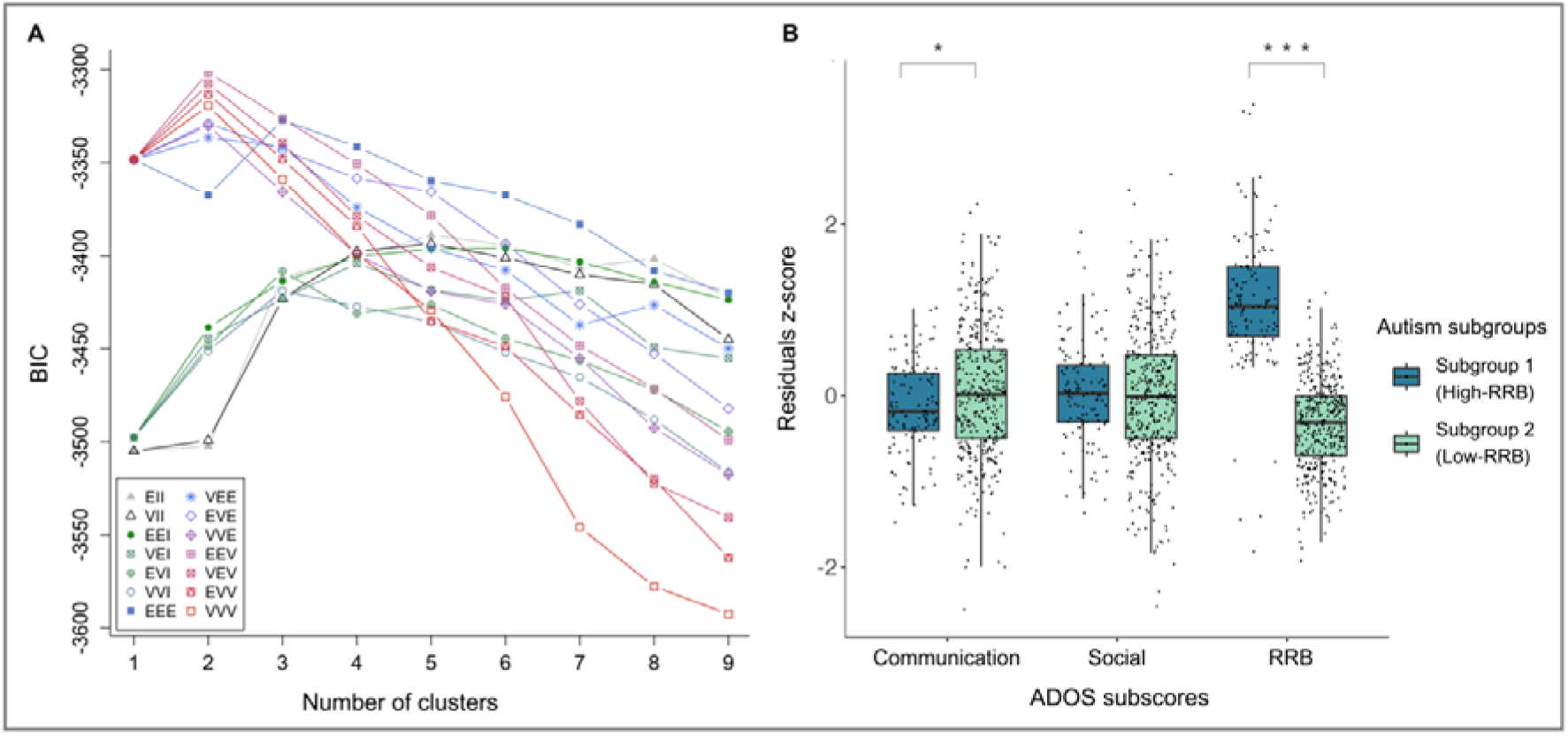
Clustering results. A) Model-based clustering, BIC. BIC = Bayesian Information Criterion. Cluster solutions between 1 and 9 are shown. Each line represents a possible model used in the model-based clustering and the three-letter code represents different combinations of geometric shapes for the clusters. The first letter indicates if the volume is constrained to be equal (E) or varies (V) across clusters. The second letter denotes if the shape is equal (E) or varies (V), or if the clusters are spherical (I). The third letter refers to the orientation of the clusters and can be equal (E), varied (V) or spherical (I). All combinations of these geographic shapes are tested (see SI). B) Differences in subscores of the ADOS-G between the two subgroups identified by model-based clustering. Subgroup 1 showed significantly higher RRBs, while subgroup 2 demonstrated significantly higher communication scores.

**Table 2.**
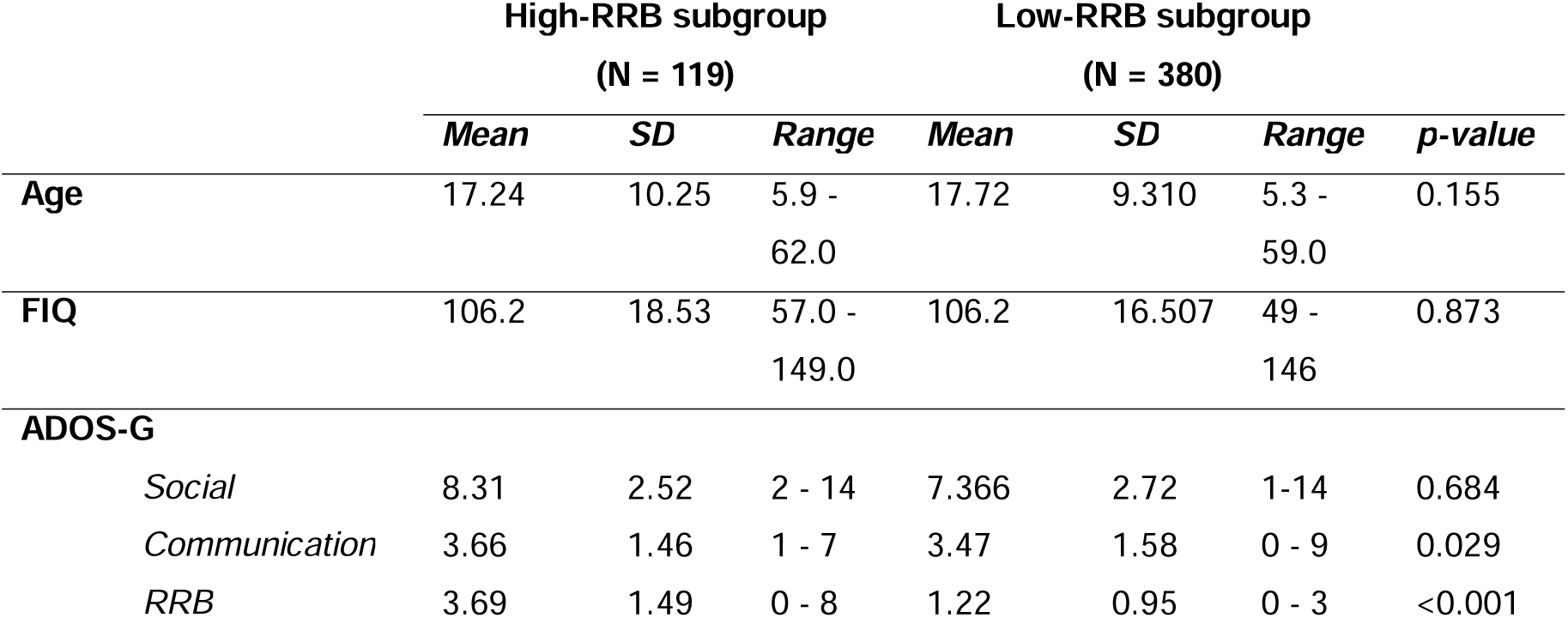
Demographics subgroups. ADOS-G = Autism Diagnostic Observation Schedule Generic. RRB = restrictive and repetitive behaviours.

|  | High-RRB subgroup<br>(N = 119) |  |  | Low-RRB subgroup<br>(N = 380) |  |  | *p-value* |
| --- | --- | --- | --- | --- | --- | --- | --- |
|  | *Mean* | *SD* | *Range* | *Mean* | *SD* | *Range* |  |
| **Age** | 17.24 | 10.25 | 5.9 -<br>62.0 | 17.72 | 9.310 | 5.3 -<br>59.0 | 0.155 |
| **FIQ** | 106.2 | 18.53 | 57.0 -<br>149.0 | 106.2 | 16.507 | 49 -<br>146 | 0.873 |
| **ADOS-G** |  |  |  |  |  |  |  |
| *Social* | 8.31 | 2.52 | 2 - 14 | 7.366 | 2.72 | 1-14 | 0.684 |
| *Communication* | 3.66 | 1.46 | 1 - 7 | 3.47 | 1.58 | 0 - 9 | 0.029 |
| *RRB* | 3.69 | 1.49 | 0 - 8 | 1.22 | 0.95 | 0 - 3 | <0.001 |

### Neuroanatomical heterogeneity and homogeneity

The neurobiological deviations obtained from the normative model were highly heterogeneous in autistic individuals, with some individuals primarily displaying positive deviations and others primarily displaying negative deviations. This can be observed in the individual deviation maps of the top 10 deviating autistic individuals, as shown in Figure S5.

The analysis whether subgrouping increases biological homogeneity in autism revealed for positive deviations (i.e., thicker CT) that survived FDR-correction that the high-RRB subgroup showed greater overlap in bilateral cerebellum and subcallosal cortex, while the low-RRB subgroup showed greater overlap in the left cerebellum. For negative deviations (i.e., thinner CT, lower volume) that survived FDR-correction, only the low-RRB subgroups showed greater overlap in left postcentral gyrus, bilateral occipital fusiform cortex, right occipital pole, left amygdala and left hippocampus than the entire autism group (see Figure 2). These results were confirmed by the sensitivity analyses using FDR-correction of the NPMs (see Supplementary Results and Figures S6).

**Figure 2.**
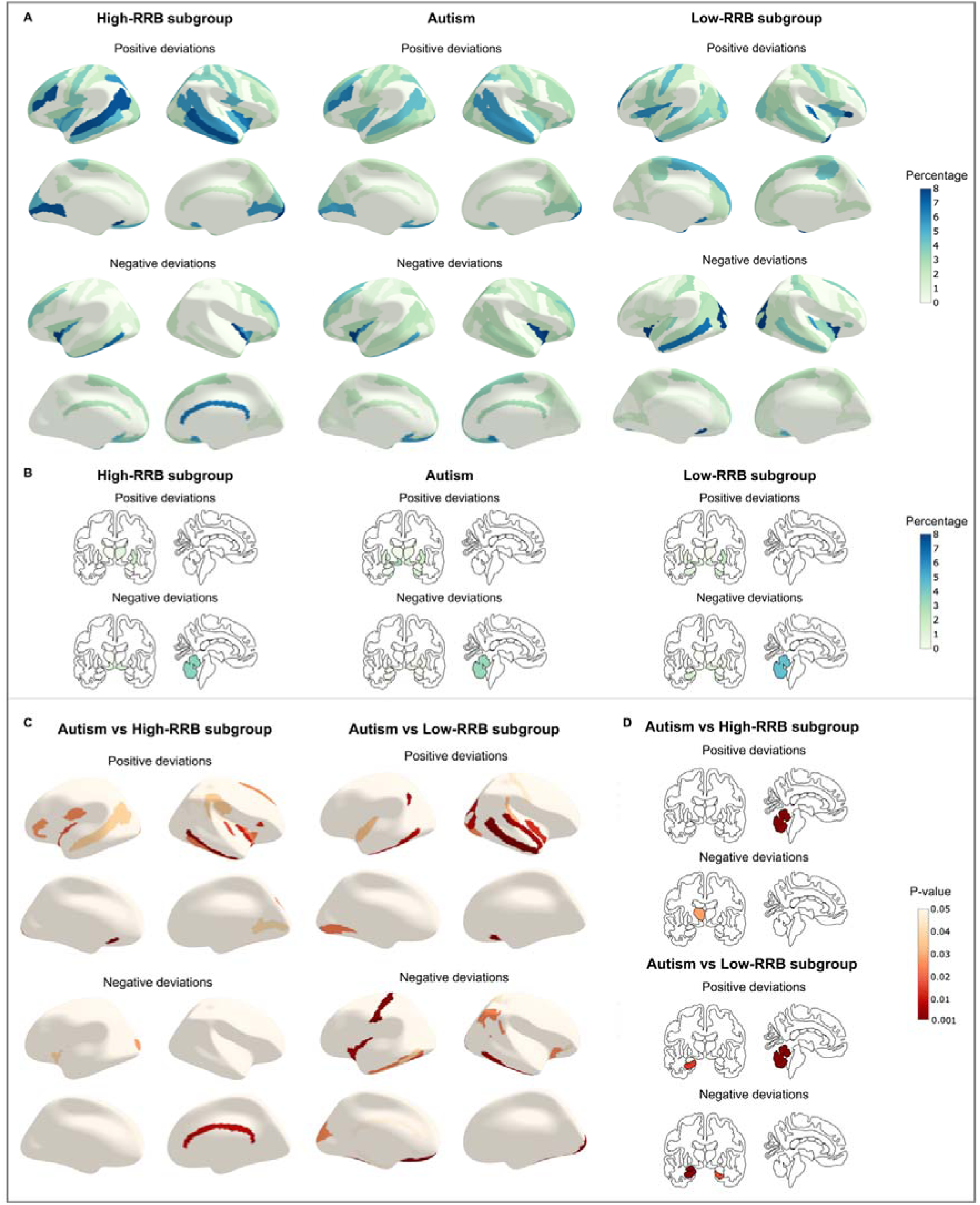
Overlap in deviation scores. A) Overlap in extreme (|Z|>2.6) cortical thickness deviations in percentage. B) Overlap in extreme subcortical volume deviations in percentage. C) Uncorrected p-values from permutation-based comparisons of overlap in cortical thickness between the subgroups and the entire autism group. D) Uncorrected p-values from permutation-based comparisons of overlap in subcortical volume between the subgroups and the entire autism group.

Finally, SVM classification between the entire autism group and NAI showed a balanced accuracy of 65%. The classification did not improve after subgrouping, with a balanced accuracy of 66% for the high-RRB subgroup and 65% for the low-RRB subgroup. Table S3 presents all other evaluation metrics.

## Discussion

In this study, we aimed to identify clinically meaningful subgroups in autistic individuals and characterize their unique neuroanatomical profiles. Our analyses revealed two robust subgroups most strongly differentiated by differences in RRBs. While we observed overall large heterogeneity in CT and subcortical volume deviations, the neuroanatomical profiles within the clinical subgroups presented more homogenous compared to the entire autism group. This work highlights the importance of parsing heterogeneity at the clinical level to help inform neurobiological underpinnings of differential symptom profiles. In the following, we incorporate these findings into the current theories and previous findings about clinical subgrouping and neurobiological heterogeneity in autism.

Prior research designed to delineate clinically distinct profiles in autism has identified different clusters to ours. These studies used different diagnostic instruments or a combination of several instruments and phenotypic measures to derive clinical subgroups. These were most commonly based on diagnostic symptoms (Cholemkery et al., 2016; Hu & Steinberg, 2009; Hus et al., 2007; Lefort-Besnard et al., 2020), but also adaptive functioning (Bitsika et al., 2008; Mandelli et al., 2023), cognitive functioning (Feczko et al., 2019; Stevens et al., 2000) and language skills (Rapin et al., 2009). Studies using the ADI-R and ADOS, for example, identified subgroups that were mostly differentiated by severity across all three clinical sub-domains, rather than one as in our case (Cholemkery et al., 2016; Hu & Steinberg, 2009; Lefort-Besnard et al., 2020) and associated with core demographic variables (Bitsika et al., 2008; Lefort-Besnard et al., 2020). In comparison, here, we *a)* used one core diagnostic measure (i.e., ADOS-G) which was widely available across a large number of individuals to circumvent the common problem of limited power and *b)* made sure to account for core demographic measures before clustering analyses to obtain clinical subgroups that do not co-segregate with other phenotypic features. Also, previous studies used different clustering methods, whereas here, we decided to use a robust model-based clustering algorithm which is flexible to accommodate clusters with varying shapes, sizes, and orientations while also providing soft probabilistic assignments to clusters. Thus, not only population heterogeneity, but also heterogeneity in methods may have resulted in different subgroups across studies.

Here, the two derived subgroups suggest that regardless of basic demographic information, autistic individuals in the ABIDE datasets present as a heterogenous group particularly with regards to RRBs. This is in line with accounts of autistic individuals varying considerably in their core clinical features ranging between mild, moderate, and severe symptoms. Particularly, RRBs are a complex category comprising both lower-order motor behaviours and higher-order cognitive behaviours (Turner, 1999). Previous studies have shown that RRBs can have adverse effects such as increased anxiety, stress, difficulties in acquiring skills, reduced social interactions, and, in some cases, self-injurious behaviours (Chaxiong et al., 2022; Jaffey & Ashwin, 2022; Rodgers et al., 2012; Rodriguez et al., 2012; Sellick et al., 2021). Behavioural programmes specifically targeting RRBs are still most effective and safe in reducing RRBs (Kodak & Bergmann, 2020) and mitigating their negative impacts (Boyd et al., 2012; Lin & Koegel, 2018; Turner-Brown & Frisch, 2020; Yaari & Dissanayake, 2021). By using subgroups, interventions can be further refined to address the needs of individuals with higher RRBs specifically.

Neuroanatomically, the two clinical subgroups showed reduced regional heterogeneity in several regions compared to the entire group of autistic individuals. This is in line with our hypothesis that grouping autistic individuals by more homogenous clinical profiles also increases their similarity at the neurobiological level. Each clinical subgroup was associated with larger deviations in different sets of regions implicating a consistently implicated neural signature associated with distinct clinical features. Interestingly, the high-RRB subgroup showed increased volume deviations particularly in the cerebellum compared to the entire group of autistic individuals. While cerebellar differences in neuroanatomy have previously yielded inconsistent results in autism (Laidi et al., 2022) and associations with social functioning (Elandaloussi et al., 2023), a large body of research also points to the involvement of cerebellar neuroanatomy and neurocircuitry in the etiology of RRBs in autism (Tian et al., 2022). In line with our findings, autistic individuals with high RRBs were found to have increased GM volume in the vermis VIII and left cerebellar lobule VIII (Seng et al., 2022) and children with complex motor stereotypies have been shown to have increased GM volume in the anterior vermis (Dean et al., 2022). Such structural studies are corroborated by functional work showing for example atypical cerebellar-cortical connectivity being related to elevated RRBs in autistic individuals (Lidstone et al., 2021). Further research into the specific localized volumetric increases in cerebellum, their developmental trajectories and associated molecular mechanisms will hold important insights into the development of potential pharmacological therapeutic interventions aimed at dysfunctional RRBs.

At the same time, the second subgroup was primarily characterized by lower RRBs and higher communication difficulties and was associated with more consistent negative deviations and thus thinner cortex in postcentral gyrus, occipital and fusiform cortex and reduced subcortical volume in amygdala and hippocampus compared to the entire group of autistic individuals. In line with this, a recent study examining longitudinal CT change in autism reported an association between less RRBs and reduced cortical thickness over time in regions such as the visual cortex, postcentral gyrus and fusiform gyrus (Bieneck et al., 2021). Also, volumetric differences in the fusiform-amygdala system (Dziobek et al., 2010; Schultz, 2005) and in the hippocampus and amygdala in young autistic children have been associated with atypical social cognition (Li et al., 2023; Munson et al., 2006; Schumann et al., 2009).

While our results confirm that subgrouping increases homogeneity, we acknowledge that there remains substantial between subject heterogeneity within the clinical subgroups. This is further highlighted by the pronounced within-group between-subject heterogeneity in deviations as depicted in Figure S5 and by low biological separability as shown by the lack of our classifier to distinguish between the two subgroups based on their neuroanatomical features. Both social-communication difficulties and RRBs are broad, multifaceted categories comprising different lower- and higher-order subcomponents that we were not able to differentiate between given the limitations of the ABIDE dataset. Research shows however that different subcomponents may be associated with different brain circuit dynamics (Supekar et al., 2021). Furthermore, different genetic, environmental, and neurobiological factors and pathways can contribute and result in the manifestation of similar phenotypic expressions, known as equifinality (Cicchetti & Rogosch, 1996). This means that despite clinical homogeneity, neuroanatomic heterogeneity is still likely to exist. Ideally, future research should address this in deeply phenotyped datasets that also provide item-level scores (Llera et al., 2022). Also, while in our case the relatively small sample sizes in the two clusters prevented us from further exploring the neuroanatomical heterogeneity, future large-scale studies can further cluster the clinically identified subgroups at the neuroanatomical level as a way to address equifinality (Buch et al., 2023; Hong et al., 2020; Lombardo et al., 2019; Shan et al., 2022; Zabihi et al., 2020).

Further limitations have to be taken into account. Due to limited power, we excluded all females, however, sex-differential factors in the manifestations of autism phenotypes are increasingly being recognized (Braden et al., 2021; Floris et al., 2023). Particularly, the expression and nature of RRBs differ between autistic males and females (McFayden et al., 2020) which also extends to their neural underpinnings (Supekar & Menon, 2015) and highlights the importance of including autistic females into studies. Finally, it has to be pointed out that heterogeneity in clinical profiles extends to level of cognitive functioning, language abilities and co-occurring conditions that are common in autism such as ADHD, anxiety and OCD. Normative modeling has previously been used to characterized transdiagnostic (Parkes et al., 2021) and disorder-specific neuroanatomical features (Segal et al., 2023) and future research will have to take both core clinical and transdiagnostic aspects into account to appreciate the complex nature of autism.

Taken together, in this work we identified clinical subgroups of autistic individuals that were not obscured by basic demographic variables and were characterized by more homogenous neuroanatomical signatures. Although we could demonstrate that clinical subgrouping reduces neuroanatomical heterogeneity, simultaneously, there remained substantial heterogeneity potentially pointing towards multiple pathways resulting in high RBBs and neurobiological subgroups with similar clinical phenotypes. Differential patterns of autistic symptoms can serve as a compass for screening and diagnosis and inform future research into the etiology and developmental trajectory of RRBs and other core autistic features. RRBs specifically will require further attention to stratify autistic individuals into subgroups with differential needs which has the potential to finding therapeutic targets and individualised support.

## Author contributions

- **Conceptualization**: NL, DLF, JM
- **Data curation**: JM
- **Formal analysis**: JM, NL, BHV
- **Funding acquisition**: NL, DLF
- **Investigation**: JM
- **Methodology**: NL, DLF, JM, BHV, ZBT, CE
- **Project administration**: NL, DLF, JM
- **Resources**: NL
- **Software**: JM, NL, DLF, BHV, CE, ZBT
- **Supervision**: NL, DLF
- **Validation**: JM, NL, DLF, BHV
- **Visualization**: JM
- **Writing—original draft**: JM, DLF, NL
- **Writing—review and editing**: JM, DLF, NL, BHV, CE, ZBT

## Data Availability

This study made use of the publicly available ABIDE datasets. This data is available at https://fcon_1000.projects.nitrc.org/indi/abide/

## Acknowledgments

The authors thank all investigators and contributors to the Autism Brain Imaging Data Exchange for their efforts in data collection and sharing and all participants and their families for participating in the study. DLF is supported by funding from the European Union’s Horizon 2020 research and innovation programme under the Marie Skłodowska-Curie grant agreement No 101025785. BHV is supported by the Swiss National Science Foundation [10001C_197480].

