## Supplementary information for "Towards Understanding Autism Heterogeneity: Identifying Clinical Subgroups and Neuroanatomical Deviations"

### **Supplementary Methods**

### Participants, study design and exclusion criteria

We combined the Autism Brain Imaging Data Exchange (ABIDE) repositories I (Di Martino et al., 2014) (<http://fcon_1000.projects.nitrc.org/indi/abide/abide_I.html>) and II (Di Martino et al., 2017) (<http://fcon_1000.projects.nitrc.org/indi/abide/abide_II.html>) for all the analyses conducted in the present study. ABIDE I includes in total 1112 participants (539 autism, 573 non-autistic individuals [NAI]) from 17 different sites, and ABIDE II includes 1114 participants (521 autism, 593 NAI) from 19 different sites, with ages ranging from 5 to 64 years. Institutional Review Board (IRB) approval to participate, or explicit waiver to provide fully anonymized data, was required prior to data contribution. All contributions were based on studies approved by local IRBs. For the autism group, autism diagnosis was determined by clinician’s consensus supported by either one or both ‘gold-standard’ diagnostic instruments, i.e., an Autism Diagnostic Observation Schedule (ADOS) (Lord et al., 2000) and/or the Autism Diagnostic Interview-Revised, (ADI-R) (Lord et al., 1994) in all sites but two (UCD and Stanford sites only used diagnostic cut-offs of ADOS and/or ADI-R for inclusion).

Sites with same/highly similar imaging acquisition protocols (as outlined and recommended on the website) were merged across ABIDE I and ABIDE II into one single site to increase sample sizes per site (i.e., KKI and ABIDEII-KKI; NYU and ABIDEII-NYU_1; SDSU and ABIDEII-SDSU; and UCLA_1 and ABIDEII-UCLA_1). Detailed information on imaging acquisition parameters can be found on the above websites.

For analyses, we excluded female participants due to scarce availability across sites (N autism=130, N NAI=280) and participants whose ADOS-Generic (ADOS-G) had not been administered by research-reliable personnel (as indicated in the phenotypic file; N autism=69). For the clustering analysis in autistic individuals (based on clinical scores), we further excluded participants with missing ADOS-G scores (N autism=348) and full-scale IQ (FIQ; N autism=55). This resulted in a sample of 499 autistic individuals who were included in the clustering analysis. For the normative modeling analysis (based on neuroimaging data), we further applied the following exclusion criteria: *a)* individuals who did not have available neuroimaging data (N autism=71, N NAI=37); *b)* participants who had low image quality as assessed by both visual inspection and the Euler number (N autism=15, N NAI=7). The Euler number is related to the number of holes in the reconstructed surface, and represents its complexity (Dale et al., 1999). In line with previous reports that also utilized ABIDE I and ABIDE II(Bethlehem et al., 2020), we excluded individuals with values smaller than -300; *c)* corrupted preprocessing, meaning e.g., participants with cortical thickness (CT) values of zero (N autism=4, N NAI=1); *d)* participants with missing FIQ values (as this was a covariate in the normative model, N autism=47, N NAI=66); and *e)* sites that contained less than ten individuals per diagnostic group to provide sufficient samples per sites and group for the training of the normative model (Bayer et al., 2022; Gaiser et al., 2023) (IP: autism N=11, NAI N=1; KUL: N autism=28; NYU_2: N autism=24; SBL autism: N=5; SDSU, N autism=1, N NAI=14). This resulted in a sample of 690 autistic and 744 non-autistic individuals between 5-64 years who were included in the normative modeling analysis. A flowchart depicting the selection process for each analytical step is visualized in Figure S1.

### Model-based clustering

To identify clinically distinct subgroups in autism, we applied model-based clustering (Banerjee & Shan, 2011) to the three subscores of the Autism Diagnostic Observation Schedule Generic (ADOS-G) (Lord et al., 2000), i.e., social interaction, communication, and restricted and repetitive behaviours (RRBs). While different ADOS-versions (i.e., ADOS-G [N=622] and ADOS-2 [N=560]) are available in ABIDE, ADOS-G has higher numbers thus not further limiting sample size.

First, we z-standardized the three ADOS-G sub-scores to account for differences in scale between them. Unlike other studies that performed clustering analyses on non-residualized clinical measures (Bitsika et al., 2008; Cholemkery et al., 2016; Georgiades et al., 2013; Hu & Steinberg, 2009; Lefort-Besnard et al., 2020; Lombardo et al., 2016), here we explicitly made sure to account for confounding variables, such as age, acquisition site, FIQ, the ADOS-G module, and the other two subscores using a General Linear Model (GLM) (see 1-3). The ADOS-G module (i.e., in this case module 2-4) indicated the specific ADOS-G test used, which differs for age groups and the expressive language level of the participant (Lord et al., 2000). The residuals from the GLMs were used for the subsequent analyses.

**1) ADOS** $\boldsymbol{social} \boldsymbol{interaction} \boldsymbol{subscore} \sim age+site+FIQ+ ADOS module+ADOS communication subscore+ADOS stereotyped behaviour subscore$

**2) ADOS** $\boldsymbol{communication} \boldsymbol{subscore} \sim age+site+IQ+ ADOS module+ADOS social interaction subscore+ ADOS stereotyped behaviour subscore$

**3) ADOS** $\boldsymbol{stereotyped} \boldsymbol{behaviour} \boldsymbol{subscore} \sim age+site+IQ+ ADOS module+ADOS social interaction subscore+ ADOS communication subscore$

Next, we employed a model-based, unsupervised data-driven clustering method (Banerjee & Shan, 2011) to identify clinical subgroups in autistic individuals. In contrast to more traditional, heuristic-based clustering algorithms (such as e.g., *k*-means clustering) with hard cluster assignments, model-based clustering provides a probability and uncertainty for each cluster. It further has the benefit of also automatically identifying the optimal number of clusters by comparing a range of different Gaussian mixture models. Each mixture model consists of a unique covariance matrix with a specific geometric interpretation of the clusters. This covariance matrix determines the volume, shape and orientation of each cluster. Here, the volume is constrained to be equal (E) or vary (V) across clusters; the shape can be equal (E) or vary (V), or the clusters can be spherical (I); the orientation of the clusters can be equal (E), vary (V) or spherical (I). All combinations of these geographic shapes are tested. The optimal number of clusters and mixture model is determined according to the Bayesian Information Criterion (BIC). Here, we used the ’mclust’ package in Rstudio (version 2022.12.0+353) (Scrucca et al., 2016) where the density estimation is based on parameterized finite Gaussian mixture models which are estimated by EM algorithm initialized by hierarchical model-based agglomerative clustering. Further, to assess the stability of the clustering results, we performed bootstrapping with 10.000 iterations using the ‘fpc’ package in Rstudio (Tian et al., 2021). Stability was assessed with the Jaccard similarity index ranging between 0 and 1. A cluster can be considered stable if its mean score is greater than 0.75 (Hennig, 2007, 2008).

Finally, we tested for differences in subscores between the identified subgroups using Wilcoxon signed-rank tests. To avoid inflation of the Type I error rate (as in this case the null hypothesis is a function of the data) (Gao et al., 2022), we additionally used the ‘clusterpval’ package in RStudio with 10.000 draws to determine the significant differences in means between the clusters (Gao et al., 2022; Winter & Hahn, 2022).

### Image processing and quality control

We processed the T1-weighted images from the ABIDE datasets using the FreeSurfer software (version 7.3.2) (Fischl, 2012). For the full pipeline and documentation, please refer to the following website: <https://surfer.nmr.mgh.harvard.edu/>. In brief, the FreeSurfer pipeline consists of intensity normalisation, skull stripping and grey (GM) and white matter (WM) surface segmentation, resulting in a tessellated representation of the GM/WM boundary. The resulting surfaces were corrected for topological abnormalities and registered to a spherical atlas based on individual cortical folding patterns. CT was defined as the shortest distance between the vertices of the GM/WM boundary and the pial surface. CT was extracted for each individual using the Destrieux parcellation scheme consisting of 74 regions in each hemisphere (Destrieux et al., 2010). Participants who had low image quality as assessed by both visual inspection and the Euler number were excluded.

To label subcortical tissue classes, we further applied the volume-based stream. After affine registration with MNI305 space, initial volumetric labeling, B1 bias field intensity correction, a high dimensional nonlinear volumetric alignment to the MNI305 atlas was performed and the volume was labeled according to the ASEG atlas (Fischl et al., 2002).

### Normative modeling

Normative modeling is a novel statistical framework that allows to compare individual neuroanatomical measures (i.e., CT and subcortical volumes) to a reference population on an individual basis (Marquand et al., 2016, 2019; Rutherford et al., 2022, 2023). This method yields coherent measures of predictive confidence in addition to point estimates, meaning we can predict both the expected CT and subcortical changes and the associated predictive uncertainty for each individual allowing us to quantify the regional deviation of CT and subcortical volume from the neurotypical range.

Here, we performed normative modelling using the PCNtoolkit (version 0.26) in Python (version 3.9.16). We used a 3-knot B-spline Bayesian Linear Regression (BLR) model with SinArcinh likelihood warping and Powell optimisation, allowing us to calculate error measures in a warped space and describe deviations under a Gaussian error distribution (Fraza et al., 2021; Rutherford et al., 2022). Included covariates in the normative model were age, site, FIQ, and the Euler number and also intracranial volume for the subcortical volume based normative modeling. The dataset was split into a training set consisting of 90% of NAI and a test set consisting of all autistic individuals and the remaining 10% of NAI. We also ensured that age, FIQ and site were evenly distributed across the training and test sets. A separate normative model was built for each of the 148 CT ROIs and 23 subcortical ROIs independently. Figure S3 depicts an example of a centile map and the model’s trajectory for the left occipital pole.

Next, we computed normative probability maps, which quantify the deviation of each participant from the normative model in each ROI. These subject-specific deviation Z-scores provide a statistical estimate of how much each individual’s true CT (subcortical) value differs from the predicted CT (subcortical) value with reference to the neurotypical pattern in each ROI. More specifically, this deviation score ($Xij$) is calculated for each subject ($i$) at each ROI ($j$) and is defined as the difference in true CT ($yij$) and predicted CT ($\hat{y}ij$) while accounting for the variance of the model ($\sigma_{nj}$) and the expected variance ($\sigma_{ij}$):

$$Zij= \frac{y_{ij}- \hat{y}_{ij}}{\sqrt{\sigma_{ij}^{2}+ \sigma_{nj}^{2}}}$$

Because we estimate a separate noise parameter for each ROI, this should accommodate regional differences in population variation (for example, the estimated variance parameter will be higher in the regions where there is greater variation across individuals). Also, the Bayesian statistical model takes various sources of uncertainty into account, automatically making inferences more conservative in regions where data are sparse.

To evaluate the model performance, we generated the standardised mean squared error (SMSE), explained variance (EV), mean standardised log loss (MSLL) and the Pearson correlation between true and predicted responses (Rho) based on the NAI from the test set, as seen in Figure S4.

### Characterizing neuroanatomical homogeneity and heterogeneity

To further characterize the output from the normative model, normative probability maps (NPM) were thresholded at an absolute value of Z>|2.6| (i.e., *P* <0.005) (Floris et al., 2021; Segal et al., 2023; Wolfers et al., 2018, 2020). Based on this fixed threshold, we defined extreme positive and extreme negative deviations for each participant. Here, a positive deviation score indicated higher CT (subcortical) values compared to the reference model, and thus, a thicker cortex or increased subcortical volume. A negative deviation score indicated lower CT (subcortical) values compared to the reference model, and thus, a thinner cortex or reduced subcortical volume. To illustrate the heterogeneity in the entire autism group and the two clinical subgroups, we visualized the individual NPMs for the top ten participants with the highest deviation scores (see Figure S5). To assess whether clinical subgrouping has increased neuroanatomical homogeneity in autism, we generated spatial overlap maps separately for positive and negative deviations. This was done by summing up the number of extreme deviations in each ROI for each individual which were then divided by the total number of individuals (multiplied by 100) using the ‘nilearn’ package in Python for cortical thickness [Nilearn version: 0.10.1; Python version: 3.9.16] (Abraham et al., 2014) and the ggseg package for subcortical volume in Rstudio (Mowinckel & Vidal-Piñeiro, 2019). These spatial overlap maps represent the percentage of extreme positive and negative deviations per ROI (Floris et al., 2021; Wolfers et al., 2018). Regions with highest overlap depict where participants within each group (i.e., NAI, entire autism group, clinical subgroups) share common deviations and thus show a more consistent (homogenous) neuroanatomical signature (see Figure 2). We did this separately in the high-RRB subgroup, the low-RRB subgroup and the entire autism group which in this case was composed of the autistic individuals making up the high- and low-RRB groups and additionally those autistic individuals that had not been included in the clustering analysis (for reasons stated above). We then subtracted the entire autism group overlap map from the overlap maps of each autism subgroup to obtain overlap difference maps for each subgroup. Next, we permuted the group labels 10.000 times to derive a distribution of random overlap differences and computed for each ROI the p-values as the proportion of random values that exceed the real overlaps. Significant ROIs were corrected for multiple comparisons using FDR-correction (*p*_FDR_<0.05, one-tailed). Given our hypothesis, that we expected increased homogeneity within subgroups, one-tailed tests were applied.

### Sensitivity analysis: robustness of Z-threshold

Normative probability maps (NPM) in the main analysis were thresholded at an absolute value of Z>|2.6| (Wolfers et al., 2018) based on the following rationale: 1) a fixed threshold simplifies the comparison across individuals, which is complicated when controlling the false discovery rate (FDR) separately for each NPM; 2) FDR-correction is insensitive to an overall shift in deviations from the normative model in each subject, meaning if one subject has small deviations across the entire cortex, they may seem to have a typical pattern when using FDR-thresholding, because the overall distribution of deviations is shifted. Nevertheless, to check the robustness of our results to different thresholding thresholds, we also repeated the analyses using FDR-correction of individual NPMs.

### SVM classification

To assess the biological separability of the subgroups, we used a classification algorithm, specifically a Support Vector Machine (SVM). SVM is a supervised classification algorithm that can identify data patterns in order to assign labels to each participant (Cervantes et al., 2020; Pisner & Schnyer, 2020). We implemented the SVM using the scikit-learn package (Pedregosa et al., 2011) (version 1.2.2) in Python (version 3.10.10). The deviation Z-scores derived from the normative model for all ROIs were used as features for the classification. We used a Gaussian radial basis function as kernel, and the penalty parameter was set to the default value of 1. Further, we used class-weighting to account for the differences in sample sizes between the groups. We determined the optimal classification threshold using threshold tuning, and evaluated the classification performance using five-fold cross-validation. The SVM was first used to classify the entire autism group and NAI and next, to classify each subgroup from NAI to test whether subgrouping would increase homogeneity. Finally, we used the SVM to classify the two subgroups from each other to evaluate the biological separability.

The code used for all analyses and visualisation is available on GitHub: https://github.com/JenteMeijer1/Autism-heterogeneity

### **Supplementary Results**

#### Sensitivity analyses

The analysis using individual FDR corrections to test whether subgrouping increases biological homogeneity in autism revealed for positive deviations (i.e., thicker CT) that the high-RRB subgroup showed greater overlap in the bilateral cerebellum and subcallosal cortex, while the low-RRB subgroup showed greater overlap in the left right cerebellum. For negative deviations (i.e., thinner CT, lower volume), the high-RRB subgroup showed greater overlap in the left thalamus while the high-RRB subgroup showed greater overlap in the bilateral occipital fusiform cortex, right occipital pole, left superior occipital gyrus, left inferior temporal gyrus, left pericallosal sulcus, long and short insular gyrus, central sulcus of the insula, left amygdala, left hippocampus, left pallidum, right accumbens and bilateral caudate than the entire autism group (see Figure S6).

### **Supplementary Figures**

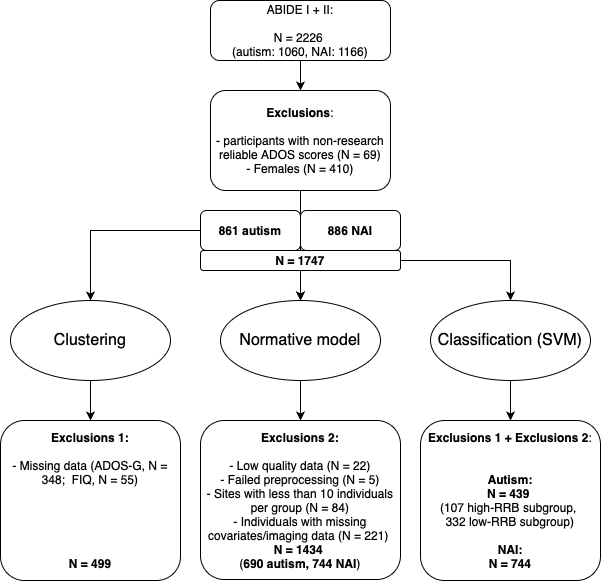

Figure S1. **Exclusions flowchart**. The ABIDE dataset consisted of 2226 participants. We excluded females because of an uneven distribution across sites, and we excluded participants from whom the clinical tests were unreliably obtained. For the clustering analysis, we used the autism group and excluded all participants with missing data for the ADOS-G and FIQ. For the normative model, we used all participants with both available MRI and covariate data, and with high-quality and successfully processed MRI scans; and sites with at least 10 participants. For the classification analyses, we applied both sets of exclusion criteria. **Abbreviations**: NAI=non-autistic individuals, FIQ=full-scale IQ.

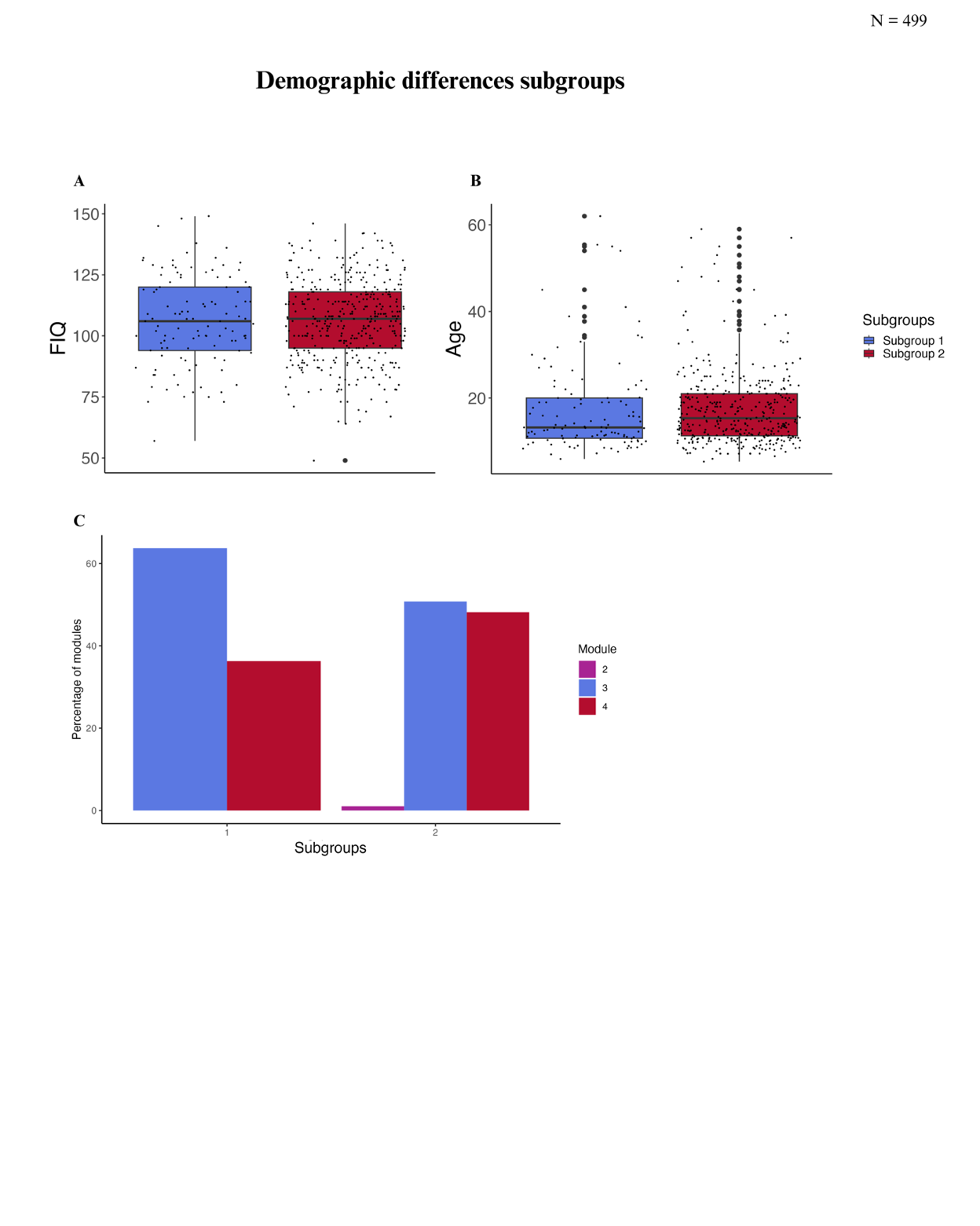

Figure S2**. Differences in demographics between clinical subgroups**. A: Full-scale Intelligence quotient (FIQ); B: Age; C: Percentage of module used. There were no significant differences in the demographics between the two clinical subgroups.

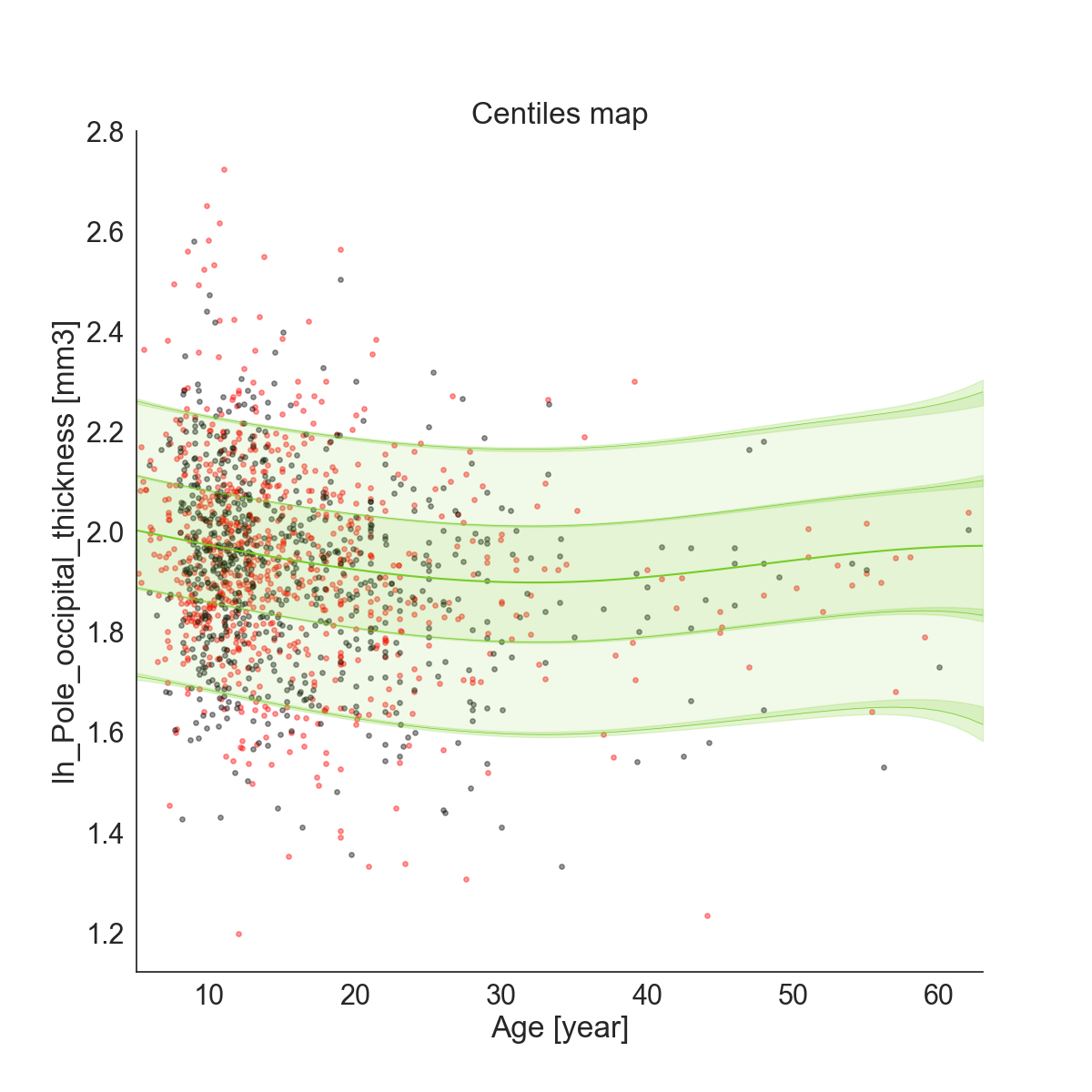

Figure S3**. Example centile map normative model**. This figure shows the centile map for the left Occipital pole, which was randomly chosen as an example. The black dots represent the train set, and the red dots the test set. The map shows the 0.25, 0.75, 0.05 and 0.95 centiles.

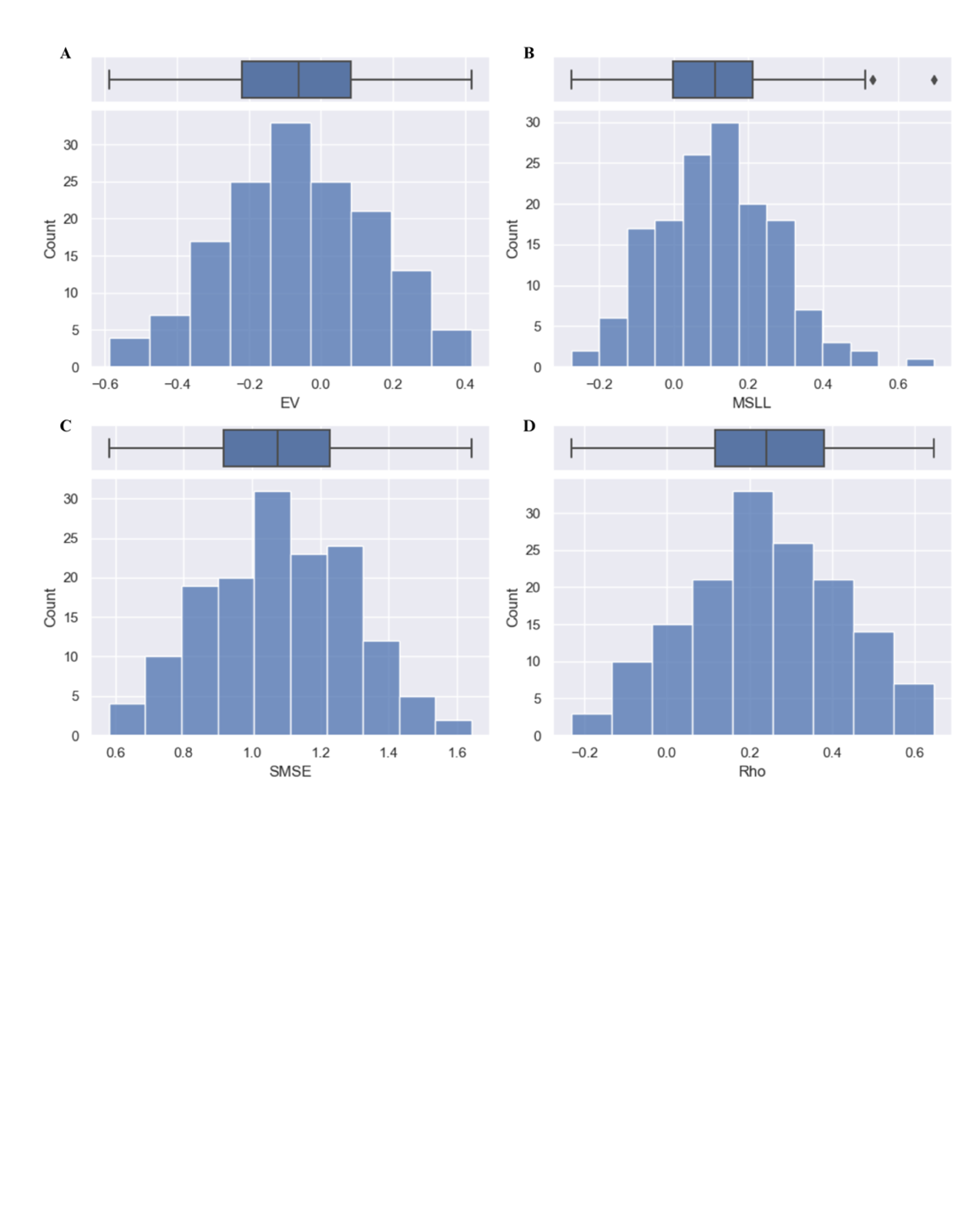

Figure S4**. Evaluation metrics normative model**. A: Explained variance (EV), closer to 1 = better model performance. B: Mean standardised log loss (MSLL), more negative = better model performance. C: Standardised mean squared error (SMSE), Closer to 0 = better model performance. D: Pearson correlation between true and predicted responses (Rho), closer to 1 = better model performance.

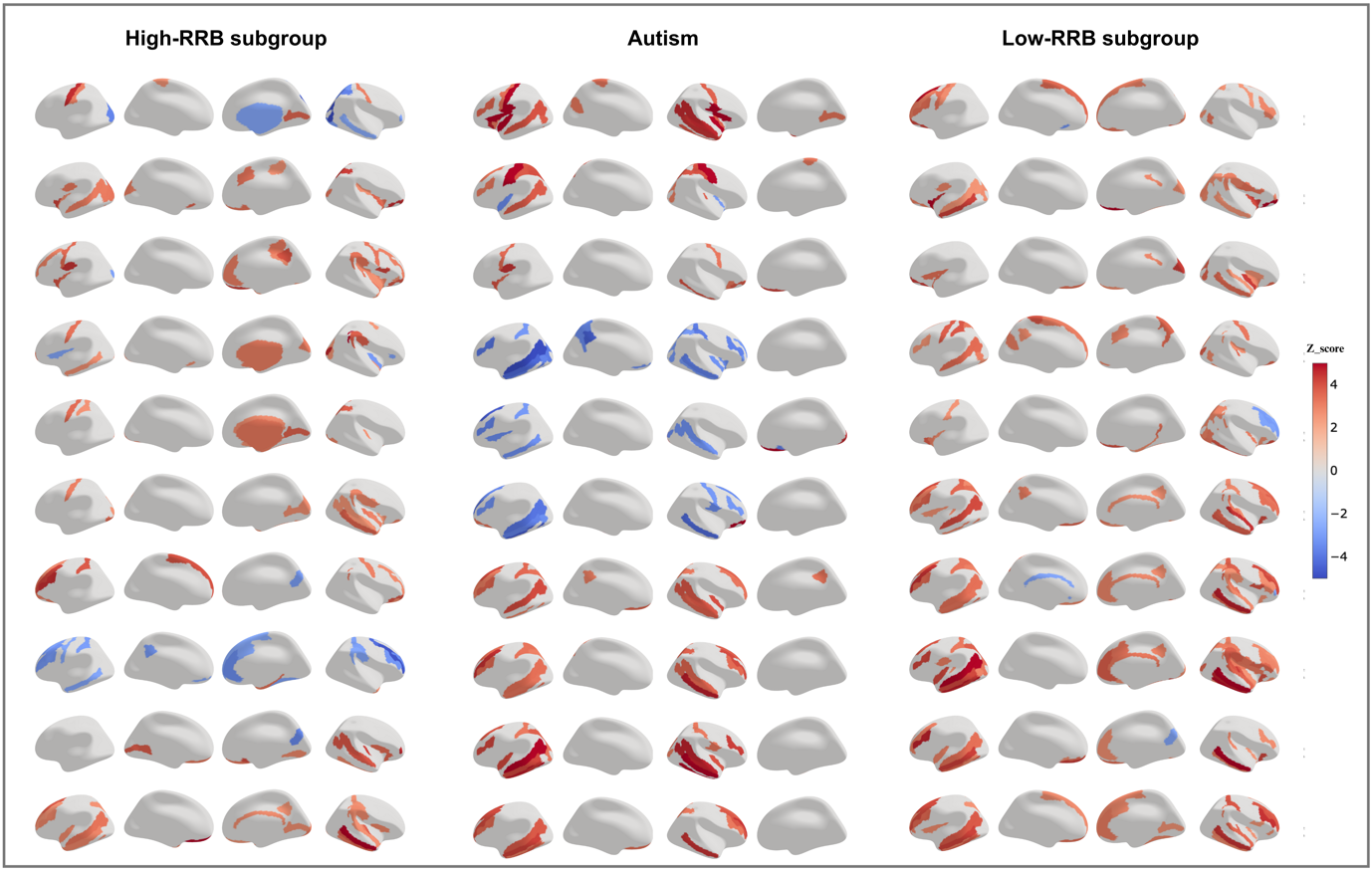

Figure S5. Individual normative probability maps for the top 10 deviating participants per group. In red positive deviations, and in blue negative deviations.

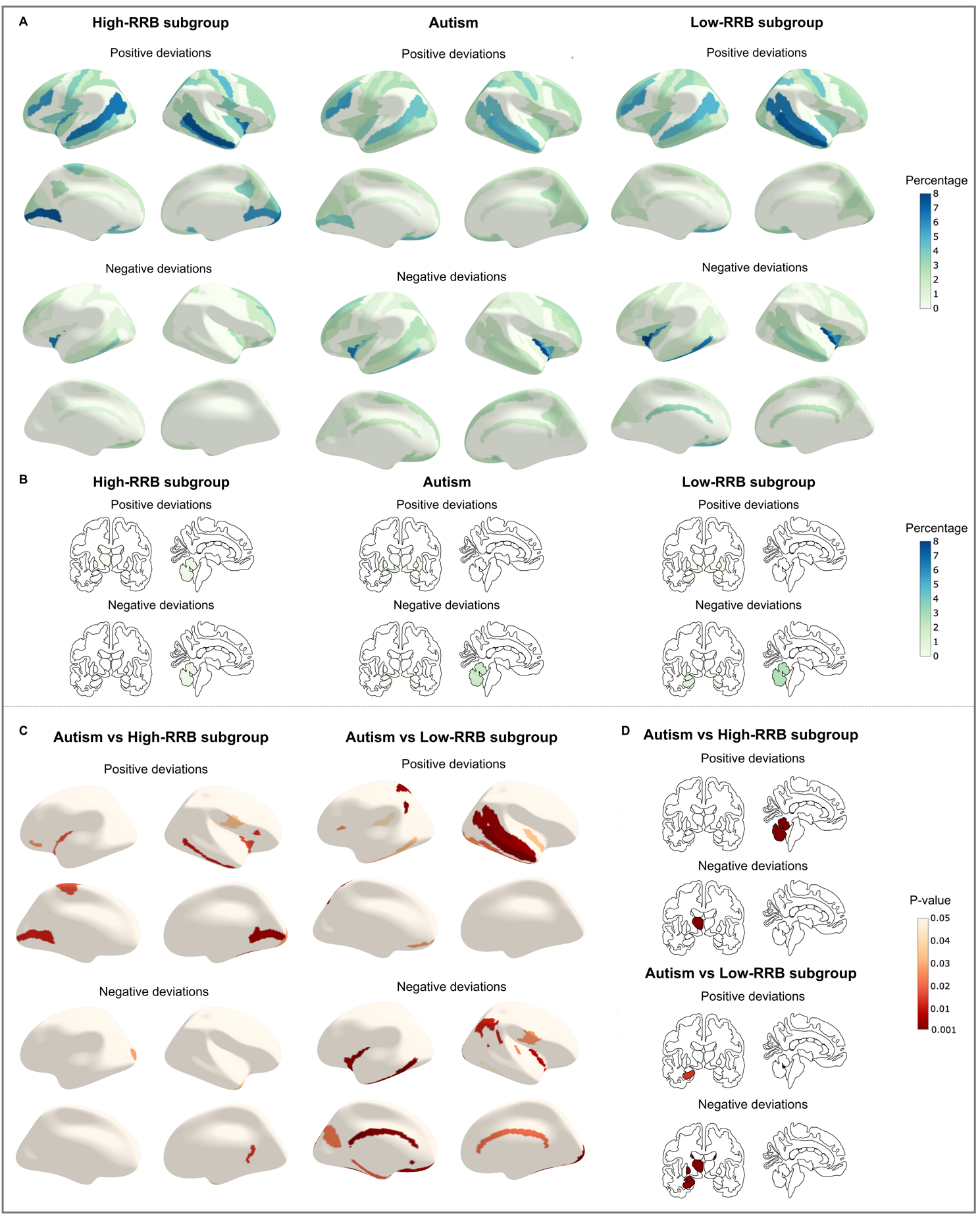

Figure S6. **Overlap in deviation scores when using individual FDR correction.** A) Overlap in extreme (individual thresholds) cortical thickness deviations in percentage. B) Overlap in extreme subcortical volume deviations in percentage. C) Uncorrected p-values from permutation-based comparisons of overlap in cortical thickness between the subgroups and the entire autism group. D) Uncorrected p-values from permutation-based comparisons of overlap in subcortical volume between the subgroups and the entire autism group.

Table S1**. Evaluation metrics for the classification**

|  | **Autism vs NAI** | **High-RRB vs NAI** | **Low-RRB vs NAI** |
| --- | --- | --- | --- |
| ***Accuracy*** | 0.61 | 0.77 | 0.60 |
| ***Balanced accuracy*** | 0.65 | 0.66 | 0.65 |
| ***ROC AUC*** | 0.70 | 0.70 | 0.69 |
